# Data Source Concordance for Infectious Disease Epidemiology

**DOI:** 10.1101/2022.06.02.22275926

**Authors:** Maimuna Majumder, Marika Mae Cusick, Sherri Rose

**Author notes:** These authors contributed equally to the manuscript and are co-first authors (Maimuna Majumder, Marika Mae Cusick).

## Abstract

**Background:** As highlighted by the COVID-19 pandemic, researchers are eager to make use of a wide variety of data sources, both government-sponsored and alternative, to characterize the epidemiology of infectious diseases. To date, few studies have investigated the strengths and limitations of sources currently being used for such research. These are critical for policy makers to understand when interpreting study findings.

**Methods:** To fill this gap in the literature, we compared infectious disease reporting for three diseases (measles, mumps, and varicella) across four different data sources: Optum (health insurance billing claims data), HealthMap (online news surveillance data), Morbidity and Mortality Weekly Reports (official government reports), and National Notifiable Disease Surveillance System (government case surveillance data). We reported the yearly number of national- and state-level disease-specific case counts and disease clusters according to each of our sources during a five-year study period (2013–2017).

**Findings:** Our study demonstrated drastic differences in reported infectious disease incidence across data sources. When compared against the other three sources of interest, Optum data showed substantially higher, implausible standardized case counts for all three diseases. Although there was some concordance in identified state-level case counts and disease clusters, all four sources identified variations in state-level reporting.

**Interpretation:** Researchers should consider data source limitations when attempting to characterize the epidemiology of infectious diseases. Some data sources, such as billing claims data, may be unsuitable for epidemiological research within the infectious disease context.

## INTRODUCTION

The COVID-19 pandemic has exposed foundational gaps in government-sponsored public health surveillance across the United States (US) (1). Most notably, for the first year of the pandemic, the Centers for Disease Control and Prevention (CDC)—which has historically been responsible for reporting population-level situational statistics (e.g., cases, hospitalizations, and deaths over time during infectious disease outbreaks)—did not efficiently report COVID-19-related statistics. This was due, in part, to lack of prioritization and underinvestment in local public health surveillance systems (2). News media organizations such as The Atlantic’s COVID Tracking Project partially filled this gap (3), highlighting the critical role that alternative data sources can play during public health emergencies. Situational statistics are also useful more broadly in infectious disease epidemiology research.

Gaps in government-sponsored public health surveillance have long preceded the pandemic, as has the practice of leveraging alternative data sources. For infectious disease research specifically, case count data obtained from news coverage of outbreaks led to studies that examined the 2014–2015 Disneyland measles outbreak (4) and the 2016 Arkansas mumps outbreak (5), as well as a broad range of international studies, including Zika (6,7) and dengue (8) in Latin America, H7N9 in China (9), and Ebola in West Africa (10), among others. News media data have repeatedly demonstrated usefulness in aggregating case counts, and in each of the aforementioned instances, were implemented to augment otherwise insufficient data from government-sponsored agencies.

In high-income settings like the US, insufficiency of government-sponsored public health data is often characterized by delays in reporting (11). Although the data may exist and are frequently treated as “ground truth” statistics, they are reported at a pace that disallows real-time evaluation of emergent crises. For example, even nationally notifiable infectious diseases are only reported once a week by the CDC’s National Notifiable Diseases Surveillance System (NNDSS) (12)—a pace that is too infrequent for real-time monitoring and mitigation of highly contagious infectious disease outbreaks. Moreover, NNDSS data—unlike news media data— are only reported at the state-level, which is an inadequate geographic resolution in the event of localized (i.e., county- or zip-level) outbreaks. The CDC also prepares Morbidity and Mortality Weekly Reports (MMWR), which include detailed government reports on notable infectious disease outbreaks (13). However, these reports are challenging to rely on for emergent crises, as there are no clear inclusion criteria for a MMWR reportable outbreak, inconsistencies in the reported level of geographic resolution, and they are often published well after the outbreak.

Beyond news media data, insurance billing claims data are also a potential alternative data source for characterizing infectious disease epidemiology in the US. These data experience more considerable delays in reporting, with data released after months or more (14,15). However, unlike the population-level situational statistics that are obtainable from news media data and government-sponsored public health surveillance systems, insurance claims provide patient-level data. Historically, these patient-level data have enabled important advances in monitoring chronic illness in both individuals and populations, but their utility within the context of acute infectious disease surveillance remains largely untested. Given recent interest in using insurance claims data to study COVID-19 (14), validating the quality of these data for other infectious diseases—those that pre-date the pandemic—is urgently needed.

In this retrospective study, we evaluate case count data for the years 2013 through 2017 from the news media platform HealthMap and the Optum insurance claims database against two government-sponsored data sources (NNDSS and MMWR) for three infectious diseases: measles, mumps, and varicella (chickenpox). Because these three diseases are nationally notifiable, healthcare providers are obligated to report cases of them to state health agencies and state health agencies are obligated to report them to the CDC—thus ensuring a high degree of completeness for government-sponsored data.

## METHODS

We compared infectious disease case counts for each disease across all four sources during the period 2013–2017. Our main outcomes of interest were yearly counts of cases and Micropolitan and Metropolitan Statistical Area (MSAs) clusters at both the national and state level. Clusters are defined as a group of cases interrelated according to both time and geography.

### Data sources

#### Health insurance claims data

Optum Clinformatics® Data Mart Database is a de-identified database derived from a large claims data warehouse(15). The claims submitted have been adjudicated to the appropriate enrollee, adjusted, and de-identified prior to inclusion in the database. Claims are subject to adjustment after initial adjudication due to delays in reporting and additional visit information.

The database includes approximately 15–20 million annual covered enrollees for a total of roughly 83 million unique enrollees from 2006–2018. During the 2013–2017 period of our study, there were approximately 39 million unique enrollees in commercial and Medicare plans. The Optum Clinformatics® Data Mart contains enrollee-level information on demographics (age and documented sex) and geography at the ZIP code level. Individual enrollee medical claims include data on the date of service, as well as associated diagnoses, procedures, laboratory tests, prescriptions, and providers.

Using a set of International Classification of Diseases (ICD-9 and ICD-10) codes, we identified enrollees with diagnoses for measles, mumps, and varicella (See Supplement Table 1 for ICD codes). Given the nature of these infectious diseases, we assumed that enrollees could only have each disease once during the five-year period. We identified service dates and ZIP codes associated with the enrollee’s first diagnosis.

US Department of Housing and Urban Development (HUD) United States Postal Service CrossWalk files were used to map patient ZIP codes to the Core-Based Statistical Areas (CBSAs) for MSAs as defined by the Office of Management and Budget (OMB) in February of 2013 (16). The Optum Clinformatics® Data Mart protects against re-identification by associating enrollee with multiple different ZIP codes if they live in a ZIP code with a small number of people. In this case, we used the first identified ZIP code-MSA pairing. Further details on cleaning and processing data from the Optum Clinformatics® Data Mart are provided in the Appendix.

Enrollees without CBSA and state-level information were not included in the cluster and state-level portion of the analysis. However, enrollees without this granular location information were included in overall national case counts.

#### Online news surveillance data

HealthMap surveillance data aggregates online informal news sources for disease outbreak monitoring and public health surveillance. Since September of 2006, HealthMap has offered free access to their automated database, and many national and international organizations have used these data for surveillance activities (17). For each HealthMap alert (e.g., news article), the database contains the disease of interest, article date, associated latitude and longitude coordinates of the location (which can be used to assign MSA or state), number of confirmed cases, and number of confirmed deaths.

We used QGIS to conduct spatial joins between the latitude and longitude coordinates associated with each HealthMap alert to MSAs and states (18). All HealthMap alerts without granular location information (e.g., only at the state-level or country-level) were removed from the analysis.

Many HealthMap alerts are duplicate entries of the same disease cluster. To avoid overestimating the number of cases reported from this source, we identified clusters within this database according to time (serial intervals) and spatial (MSA) constraints. The start and end of an MSA-level cluster was determined by two consecutive serial intervals, the time between successive cases in transmission, of zero new cases. We assumed the total number of cases associated with each MSA-level cluster was the highest number of confirmed cases reported among all associated HealthMap alerts.

#### Official government reports

MMWR contain scientific records of public health information and recommendations (13). For major disease outbreaks, the CDC will publish a conclusive MMWR, describing key information such as the date of identification, locations affected, and total number of cases. We manually reviewed all MMWR that related to measles, mumps, and varicella and extracted cluster identification dates, confirmed case counts, and corresponding MSA locations to allow comparison against the other data sources considered in our analysis. The Appendix contains detailed information on all MMWR.

#### Government case surveillance data

The CDC conducts mandatory disease reporting and surveillance for our three diseases of interest. We used data from NNDSS, which provides weekly tables of disease counts (12) The data contain the number of cases reported during the current week, as well as the number of cumulative cases reported over a given year. If there is a delay in reporting, the case will only appear as a part of the cumulative count. NNDSS only provides case counts at the state-level; a more granular geographic resolution is unavailable for public use.

### Analyses

#### Standardized national yearly case counts

For each disease, we reported source-specific national yearly case counts standardized to 100,000 persons. Optum data was standardized to the total number of eligible Optum enrollees during the years 2013–2017. Data from NNDSS, HealthMap, and MMWR were standardized to the US population as per census bureau national population estimates (19). While Optum and MMWR are not meant to capture case counts in ways that are nationally representative, values are standardized to this population for comparability across data sources.

#### National cumulative case counts

For each disease and each data source, we reported cumulative incidence of cases over the entire study period. Due to Optum data privacy requirements, we display the cumulative case count once the national case-counts are at least 16 cases for this data source.

#### State-level cases

For each disease, we reported yearly state-level case counts for Optum, NNDSS, HealthMap, and MMWR. State information was ascertained from each data source. In Optum, we translated patient ZIP code information to state-level information using the pyzipcode python module (20). We used available NNDSS state-level information directly.

Confirmed cases from each HealthMap MSA-level cluster were allocated to corresponding states. In the case of multi-state MSAs, we allocated cases to states according to the relative proportion of HealthMap alert-associated states within the cluster. Finally, based on the identified MSA location from the MMWR, we allocated cases to each state. As per Optum privacy constraints, we do not report any state-level cases counts smaller than 16 cases.

#### State-level clusters

For each disease, we reported the yearly number of MSA-level clusters in a given state according to Optum, HealthMap, and MMWR. The start and end of an MSA-level cluster was determined by two consecutive serial intervals of zero new (i.e., incident) cases. Serial interval periods differ based on the disease of interest: measles (12 days), mumps (18 days), and varicella (14 days) (21). We report MSA-level clusters with at least 16 cases due to Optum privacy constraints and then comparability across all data sources. Further details regarding cluster identification are provided in the Appendix. Because NNDSS does not provide granular geographical data beyond the state-level, we did not use this source to identify MSA-level clusters.

In the event of multi-state MSAs, we assigned the MSA-level cluster to a single state for each of the sources. In Optum, we assigned the MSA-level cluster according to the most frequent patient-level state. In HealthMap, we assigned the MSA-level cluster to the most commonly reported state among the associated HealthMap alerts. Finally, for MMWR, we assigned the MSA-level cluster to states by extracting the state from the available text information, as further specified in the Appendix.

## RESULTS

National standardized incidence for all three diseases is substantially higher for Optum data in comparison to other sources (Figure 1) and implausibly large in magnitude. Case counts from MMWR are the lowest, although this is expected as MMWR are only generated for key clusters across the US. While HealthMap reports slightly higher case counts in comparison to NNDSS for measles and mumps, there were fewer cases reported in varicella, suggesting that varicella is less “newsworthy.” Unstandardized yearly case counts are provided in Supplement Figures 2–4.

**Figure 1.**
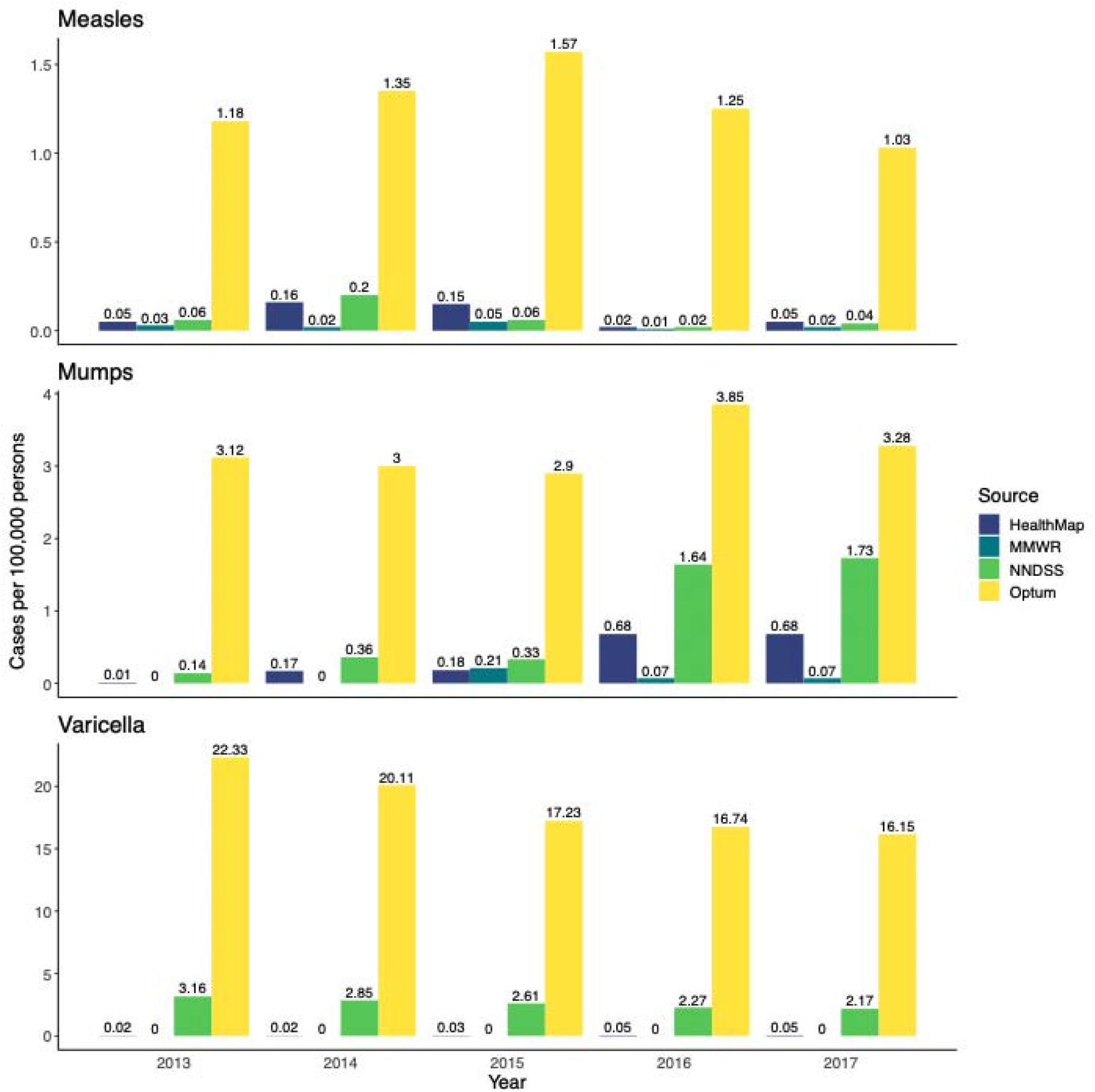
Standardized yearly national case counts *Note: MMWR and Optum are not designed to capture the entire US population; values are standardized to this population for comparability across data sources*.

Examining state-level geographic trends, Ohio had the highest number of measles cases during the study period according to HealthMap and NNDSS (Figure 2). In comparison, Optum reported the highest number of cases in New York and New Jersey. California had the highest case counts according to MMWR. All states with MMWR were also captured as having measles cases in both HealthMap and NNDSS.

**Figure 2.**
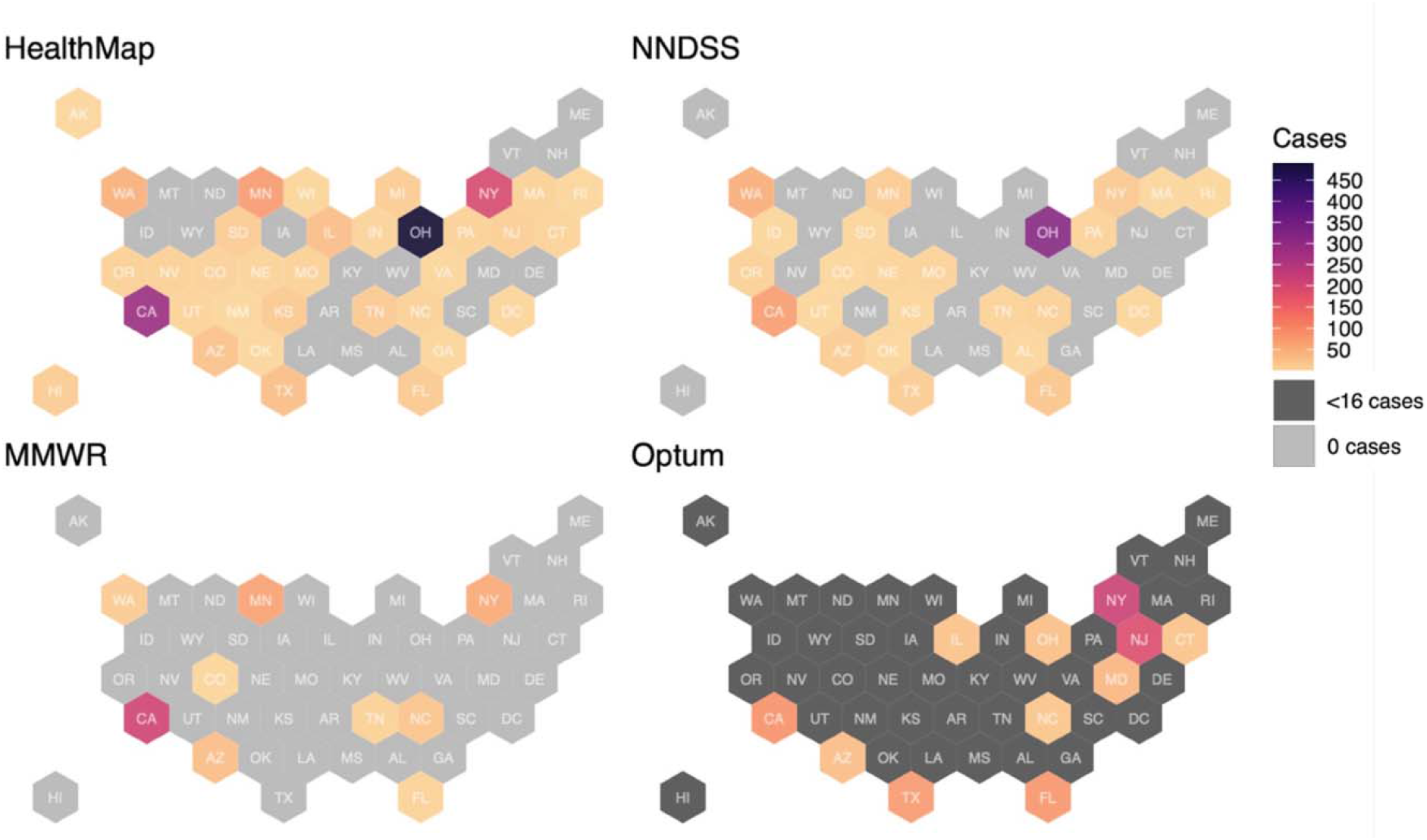
State-level measles cases (2013–2017) *Note: Optum data are presented for states with at least 16 cases during the study period*.

For mumps, there were few MMWR on outbreaks during the study period (Figure 3). Of the states with clusters identified by MMWR, all other sources reported cases for these states as well. There was a high concentration of mumps cases in the Midwestern region (Iowa, Illinois, Missouri, Indiana, and Ohio) according to HealthMap, yet this concentration was not reflected as clearly in NNDSS and Optum data. NNDSS reported a substantial number of mumps cases in Arkansas, yet there was no MMWR on these cases.

**Figure 3.**
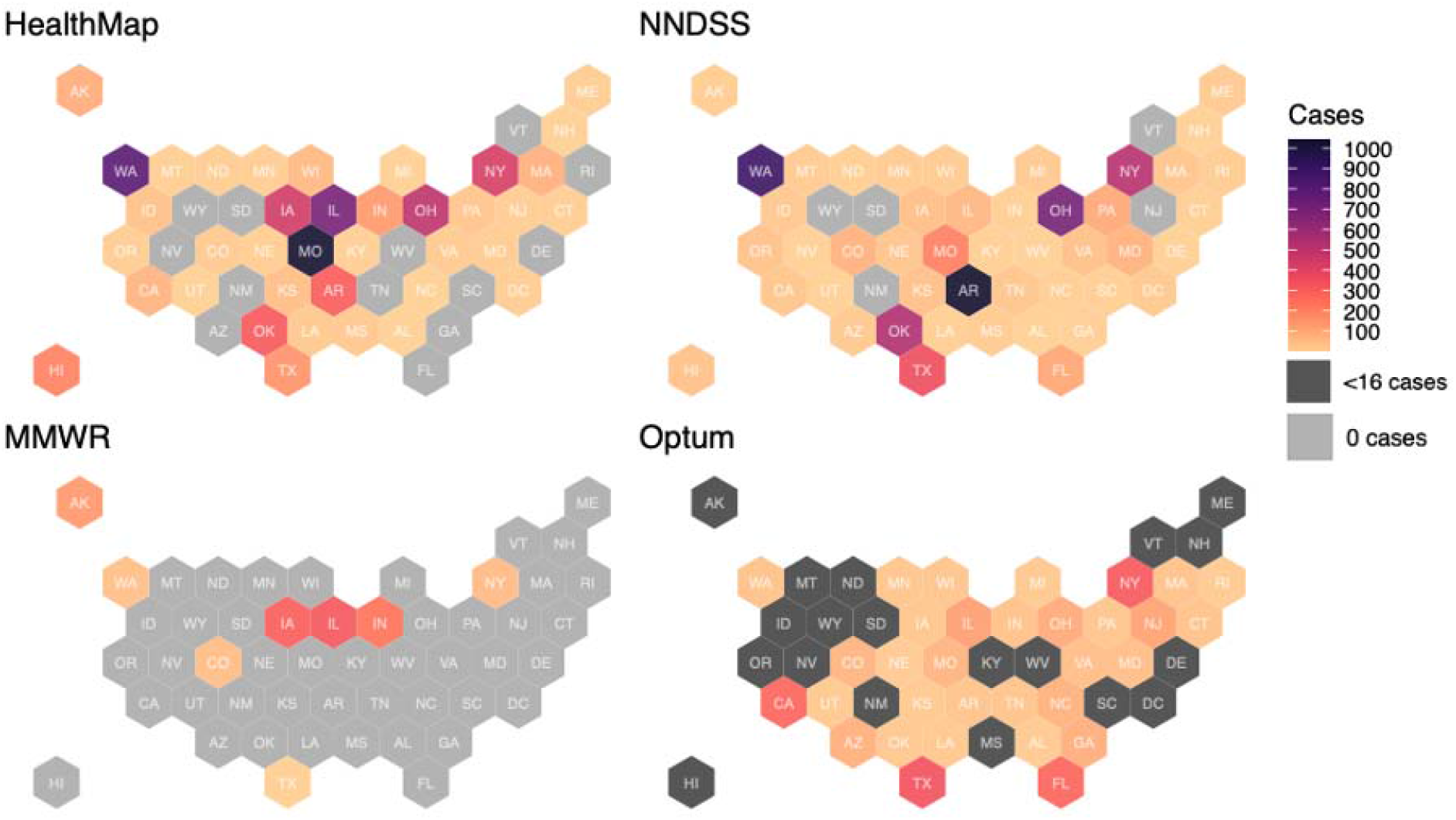
State-level mumps cases (2013–2017) *Note: Optum data are presented for states with at least 16 cases during the study period*.

Nearly all states reported varicella cases in the Optum data (Figure 4). According to NNDSS, Texas and Florida reported the highest numbers of varicella cases, which was also reflected in the Optum data, as these states also had higher numbers during the study period. Very few varicella cases were reported in HealthMap and MMWR.

**Figure 4.**
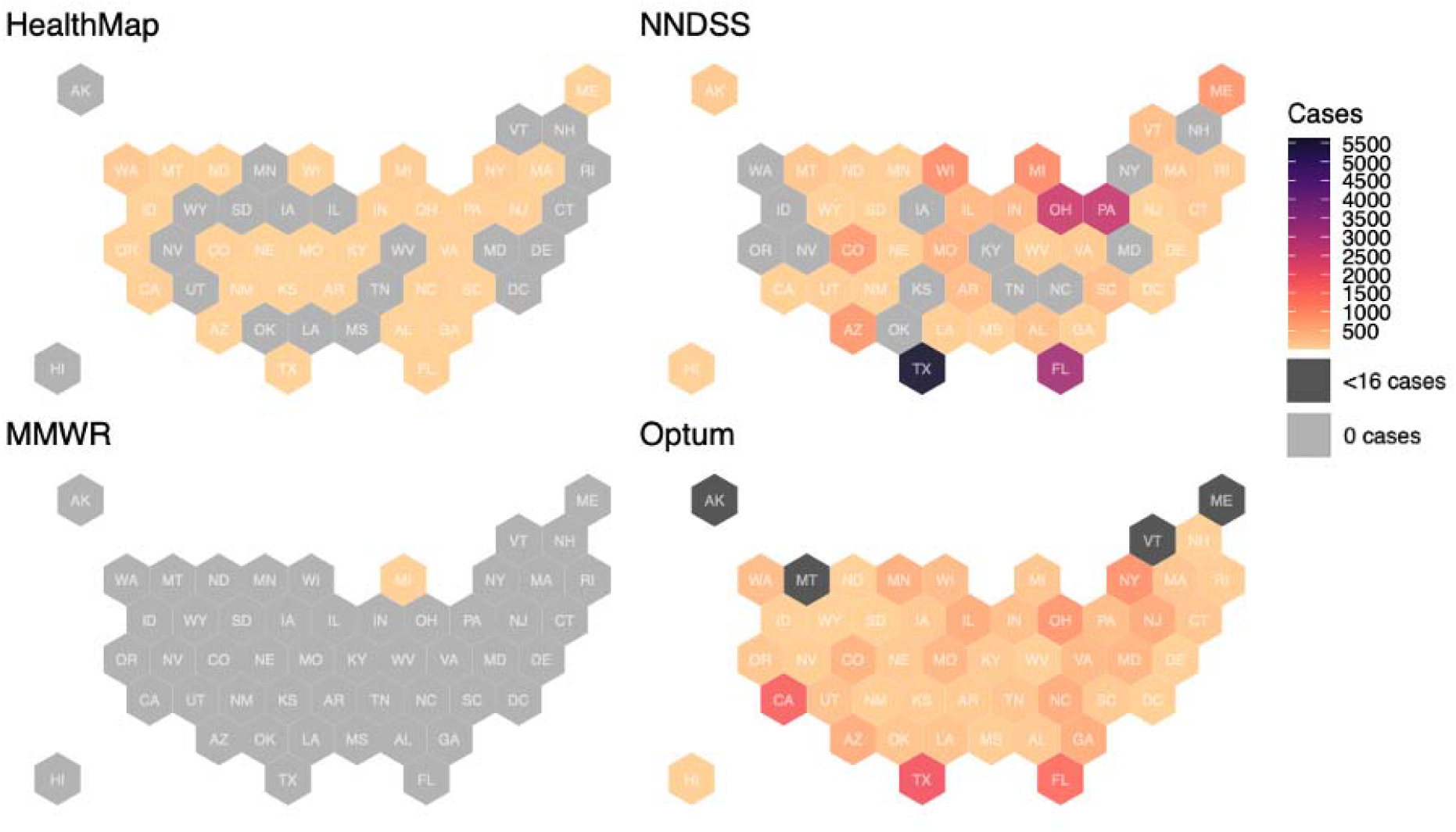
State-level varicella cases (2013–2017) *Note: Optum data are presented for states with at least 16 cases during the study period*.

**Figure 5.**
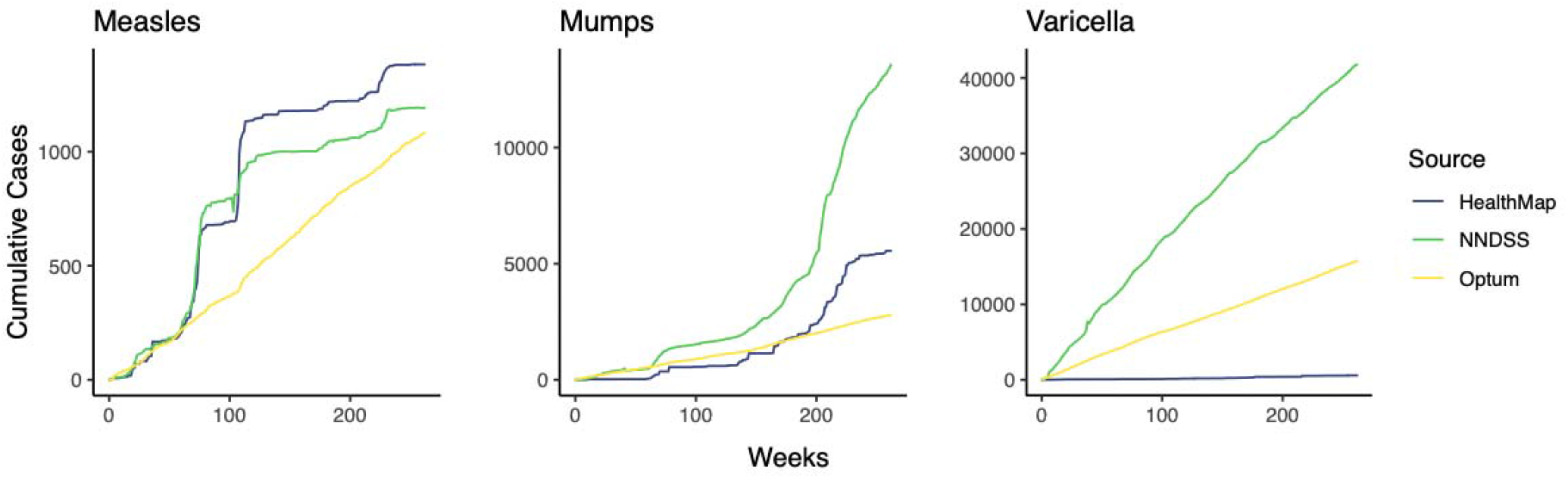
Cumulative incidence during study period

Cumulative incidence of measles and mumps cases over the study period follows similar general patterns in HealthMap and NNDSS. Disease clusters are evident as case counts rise rapidly and then are stagnant. In comparison, in the Optum data, measles and mumps case counts rose constantly over time. Incidence of varicella cases were constant over time in all data sources.

## DISCUSSION

To our knowledge, this is the first study to examine the concordance of infectious disease case counts across multiple disparate sources, including news media, insurance claims, and government-sponsored data. We found wide variation in the number of reported cases for measles, mumps, and varicella across these data sources, with implausibly high volumes of standardized cases reported by Optum that far exceed the other sources considered. Because these three highly infectious diseases are nationally notifiable and thus must be reported both to state health agencies and to the CDC, it is highly unlikely that Optum would capture cases that were not reported by NNDSS.

Overcounting may be due to the coding of likely cases, as perceived by providers, rather than laboratory confirmed diagnoses. However, laboratory results in claims data are typically incomplete as many test results are not recorded (22); thus, analyses that include only laboratory confirms cases produce severe undercounts (as presented in the Appendix.)

Notably, evidence of over-billing for conditions like measles and mumps may contribute to the rise in medical expenditures and patient healthcare spending. Using Optum data on reported total paid charges, we estimated wasted expenditures from suspected over-billing of measles and mumps cases to be roughly 395 thousand dollars for the five-year period among Optum enrollees alone. While the use of insurance claims data to characterize infectious disease epidemiology might appear appealing due to the availability of additional individual-level information, these analyses may lack credibility given the erroneous coding issues we identified here.

While there are well-known gaps in government-sponsored data sources, NNDSS compared favorably to other sources, capturing a larger scope of the mumps outbreak in 2016-17 as well as more varicella than HealthMap or MMWR. We also saw that HealthMap may produce similar case counts to NNDSS in non-outbreak years for measles and mumps. This is advantageous as HealthMap does not have the same delays in reporting that characterize NNDSS and is also available at a more granular geographic resolution. However, HealthMap is not likely to be a reliable source for case counts of less “newsworthy” diseases like varicella.

Before using a particular data source to characterize the epidemiology of a given infectious disease, researchers should consider the underlying process that led to the creation of the data and how these processes may impact reliability. Some data sources, such as health insurance billing claims, may not be suitable for research questions that are contingent on reliable reporting of situational statistics—including those that pertain to the epidemiological properties of COVID-19 and other infectious diseases.

## Data Availability

All data in the present study are available online with the exception of Optum Clinformatics® Data Mart.

https://www.cdc.gov/mmwr/index.html

https://www.cdc.gov/nndss/data-statistics/index.html

https://www.healthmap.org/en/

## DELCARATIONS

## Acknowledgements

Data for this project were accessed using the Stanford Center for Population Health Sciences Data Core. The PHS Data Core is supported by a National Institutes of Health National Center for Advancing Translational Science Clinical and Translational Science Award (UL1TR003142) and from Internal Stanford funding. The content is solely the responsibility of the authors and does not necessarily represent the official views of the NIH.

We would like to acknowledge and thank David Scales for using QGIS to do the spatial joins between HealthMap geographic coordinates and MSAs.

## Authors’ contributions

MM and SR conceptualized the research question and methodology. MC curated the data and conducted the analysis. All authors contributed to the final draft in writing, reviewing, and editing. MM and MC directly accessed and verified the underlying HealthMap, MMWR, and NNDSS data reported in the manuscript and MC and SR directly accessed and verified the Optum data due to data use agreements.

## Ethics committee approval

Access to Optum Clinformatics® Data Mart was approved by Stanford University’s Institutional Review Board (IRB) under protocol 40974.

