## Appendix for "Data Source Concordance for Infectious Disease Epidemiology"

Supplement Table 1. ICD-9/10 codes used in our analysis

| Disease | ICD-9 Codes | ICD-10 Codes |
| --- | --- | --- |
| Measles | 55.0, 55.1, 55.2, 55.9, 55.71, 55.79 | B05.4, B05.89, B05.9, B05.1, B05.0, B05.81, B05.3, B05.2 |
| Mumps | 72.0, 72.1, 72.2, 72.3, 72.9, 72.71, 72.71, 72.79 | B26.85, B26.81, B26.0, B26.3, B26.84, B26.9, B26.1, B26.2, B26.82, B26.83 |
| Varicella | 52.0, 52.1, 52.7, 52.8, 52.9 | B01.0, B01.11, B01.2, B01.81, B01.89, B01.9 |

*Optum Data Pre-Processing Procedure*

After identifying all enrollees with measles, mumps, and varicella diagnoses during the 2013–2017 period, we obtained enrollee-specific data—including the enrollee’s year of birth, gender, 5-digit ZIP code, insurance enrollment dates, and plan type—associated with the patient’s first diagnosis of each disease. A small number of enrollees in the data had multiple diagnoses for the same condition (less than 3%), but we made the assumption that each patient was only infected one time.

We used the HUD US Postal Service CrossWalk files to map the ZIP code to CBSAs and MSAs as defined by the OMB in 2013. We ensured that the mapping was eligible during the enrollee’s service date. Less than 4% of the CBSA codes obtained in the HUD mapping file were not available in the OMB 2013 definitions. Enrollees without these codes were not included in the cluster identification procedure.

*OptumInsight Clinformatics Data Mart Cohort Demographic Information*

Demographic information from the disease cohorts identified by Optum demonstrated that the cohorts skewed more heavily female, with the highest proportion in the measles cohort (Supplement Table 2). The varicella cohort was the youngest of the three cohorts, with an average age of 32, and the mode being age 1. Most enrollees across the cohorts had a commercial insurance plan rather than Medicare. Mumps and varicella cases were identified in all 50 states, and measles cases were identified in 46.

Supplement Table 2. Optum Enrollee Disease Cohort Demographics

|  |  | Measles (N=1,129) | Mumps (N=2,894) | Varicella (N=16,308) |
| --- | --- | --- | --- | --- |
| Documented Sex | Female | 645 (57%) | 1571 (54%) | 8979 (55%) |
| Age  *Mean (SD)* |  | 44 (22) | 46 (24) | 32 (27) |
| Plan type | Commercial | 819 (73%) | 1997 (69%) | 13463 (83%) |
|  | Medicare | 310 (27%) | 897 (31%) | 2845 (17%) |
| States |  | 46 | 50 | 50 |

*MSA-level Cluster Identification Procedure*

The start and end of an MSA-level cluster was determined by two consecutive serial intervals of zero incident cases. Serial interval periods differ according to the infectious disease of interest. We identified MSA-level clusters for Optum, HealthMap, and MMWR using the following procedure.

For cases with identifiable MSA information, we ordered patient cases based on the date of identification/diagnosis. For each identified case in the MSA, we checked whether the case’s date of identification/diagnosis fell between the start and end date of any prior MSA-level clusters, and thus was already part of a cluster. If the case was not already part of a cluster, we consider this identification/diagnosis date as the start of a new cluster, and we identified all individuals within 2 consecutive serial intervals of the first identified case. If there were no cases within this window, we considered 2 serial intervals after the first identified case to be the end of the cluster. If there were additional cases within this window, we identified additional cases within 2 consecutive serial intervals of the most recent date. We continued this process until there were no additional cases remaining, thus marking the end of the MSA-level cluster. As mentioned prior, we did not use NNDSS to identify MSA-level clusters because it does not contain geographical data more granular than the state-level.

*Investigation of Optum Laboratory Data*

To further decipher the large number of cases reported in Optum across all three diseases of interest, we investigated the presence of laboratory results in the data. Because the database only includes laboratory tests within certain laboratory networks, not all Optum enrollees have laboratory data. After identifying the CPT codes associated with serologic testing for each disease, we found that roughly 11% (133) of the enrollees with measles diagnoses had a measles serologic testing CPT code (86765), 15% (449) of the enrollees with mumps diagnosis had a mumps serologic testing CPT code (86735), and 11% (1768) of the enrollees with varicella diagnosis had varicella serologic testing (86787). There was substantial missingness in the laboratory data results, suggesting that the use of laboratory confirmed cases alone would not improve the reliability of the Optum data source for epidemiological research within the infectious disease setting. Laboratory data also had substantial missingness and variations in the reporting of test results, and thus, ascertaining accurate summary statistics for positive test results is not straightforward.

*MMWR Manual Curation Summaries (1)*

Measles

- Measles Outbreak in an Unvaccinated Family and a Possibly Associated International Traveler — Orange County, Florida, December 2012–January 2013
  - Date: 1/11/2013
  - Location: Orange County, Florida
  - State: Florida
  - CBSA Code: 36740
  - CBSA Title: Orlando-Kissimmee-Sanford, FL
- Notes from the Field: Measles Outbreak Among Members of a Religious Community — Brooklyn, New York, March–June 2013
  - Date: 3/13/2013
  - Location: Brooklyn, New York
  - State: New York
  - CBSA Code: 35620
  - CBSA Title: New York-Newark-Jersey City, NY-NJ-PA
- Notes from the Field: Measles Outbreak Associated with a Traveler Returning from India — North Carolina, April–May 2013
  - Date: 4/13/2013
  - Location:
    - International flight in North Carolina from India
    - Airport information was not provided
  - State: North Carolina
  - CBSA Code: N/A
  - CBSA Title: N/A
- Notes from the Field: Measles Transmission in an International Airport at a Domestic Terminal Gate — April–May 2014
  - Date: 4/22/14
  - Location: Minneapolis, Minnesota
    - Text justification: On April 17th, a child developed a measles-like rash while on an international flight from India to the United States before taking a connecting flight from Chicago to Minneapolis. On May 5th, the Massachusetts Department of Health contacted the Minnesota Department of Health to report a case of measles in a 46-year-old Minnesota resident.
  - State: Minnesota
  - CBSA Code: 33460
  - CBSA Title: Minneapolis-St. Paul-Bloomington, MN-WI
- Notes from the Field: Measles in a Micronesian Community — King County, Washington, 2014
  - Date: 5/30/14
  - Location: King’s County, Washington
  - State: Washington
  - CBSA Code: 42660
  - CBSA Title: Seattle-Tacoma-Bellevue, WA
- Notes from the Field: Measles — California, January 1–April 18, 2014
  - Date: 1/1/14
  - Location: California
    - No specific regions given
  - State: California
  - CBSA Code: N/A
  - CBSA Title: N/A
- Measles Outbreak — California, December 2014–February 2015
  - Date: 1/5/15
  - Location: Disney Theme Park (Anaheim, California)
  - State: California
  - CBSA Code: 31080
  - CBSA Title: Los Angeles-Long Beach-Anaheim, CA
- Notes from the Field: Measles Outbreak at a United States Immigration and Customs Enforcement Facility ― Arizona, May–June 2016
  - Date: 5/25/16
  - Location: Eloy ICE detention center (Pinal county), Arizona
  - State: Arizona
  - CBSA Code: 38060
  - CBSA Title: Phoenix-Mesa-Scottsdale, AZ
- Measles Outbreak of Unknown Source — Shelby County, Tennessee, April–May 2016
  - Date: 4/5/16
  - Location: Shelby County, Tennessee
  - State: Tennessee
  - CBSA Code: 32820
  - CBSA Title: Memphis, TN-MS-AR
- Measles Outbreak — Minnesota April–May 2017
  - Date: 4/10/17
  - Location: Hennepin, Ramsey, LeSueur, and Crow Wing, Minnesota
  - State: Minnesota
  - CBSA Code: 33460
    - 3 out of the four counties are in this CBSA. Most of the cases were in Hennepin
  - CBSA Title: Minneapolis-St. Paul-Bloomington, MN-WI
- Public Health Economic Burden Associated with Two Single Measles Case Investigations — Colorado, 2016–2017
  - Date: 7/7/2016
  - Location: Denver, Colorado
  - State: Colorado
  - CBSA Code: 19740
  - CBSA Title: Denver-Aurora-Lakewood, CO
- Measles outbreak associated with adopted children from China--Missouri, Minnesota, and Washington, July 2013
  - Date: 1/5/2013
  - Location:
    - Two cases of measles in recently adopted children from an orphanage in China. Both ill children traveled on different flights to the United States. They were hospitalized shortly after arrival in Washington and Missouri. Two additional cases were identified in Minnesota (another child adopted from China and an adoptive mother)
  - State: N/A
  - CBSA Code: N/A
  - CBSA Title: N/A
- Notes from the Field: Measles in a Patient with Presumed Immunity — Los Angeles County, 2015
  - Date: 2/4/2015
  - Location: Los Angeles County, California
  - State: California
  - CBSA Code: 31080
  - CBSA Title: Los Angeles-Long Beach-Anaheim, CA
- Notes from the Field: Lack of Measles Transmission to Susceptible Contacts from a Health Care Worker with Probable Secondary Vaccine Failure — Maricopa County, Arizona, 2015
  - Date:1/23/2015
  - Location: Maricopa County, Arizona
  - State: Arizona
  - CBSA Code: 38060
  - CBSA Title: Phoenix-Mesa-Scottsdale, AZ

Mumps

- Notes from the Field: Complications of mumps during a university outbreak among students who had received 2 doses of Measles-Mumps-Rubella vaccine - Iowa, July 2015-2016
  - Date: 7/1/15
  - Location: University of Iowa (Johnson Country)
  - State: Iowa
  - CBSA Code: 26980
  - CBSA Title: Iowa City, IA
- Mumps Outbreak at a University and Recommendation for a Third Dose of Measles-Mumps-Rubella Vaccine — Illinois, 2015–2016
  - Date: 5/1/15
  - Location: University of Illinois at Urbana-Champaign
  - State: Illinois
  - CBSA Code: 16580
  - CBSA Title: Champaign-Urbana, IL
- Notes from the Field: Absence of Asymptomatic Mumps Virus Shedding Among Vaccinated College Students During a Mumps Outbreak — Washington, February–June 2017
  - Date: 2/8/17
  - Location: University of Washington, Seattle
  - State: Washington
  - CBSA Code: 42660
  - CBSA Title: Seattle-Tacoma-Bellevue, WA
- Mumps Outbreak at Four Universities - Indiana, 2016
  - Date: 2/1/16
  - Location: Universities within 65 miles of Indianapolis
  - State: Indiana
  - CBSA Code: 26900
  - CBSA Title: Indianapolis-Carmel-Anderson, IN
- Mumps Outbreak in a Marshallese Community — Denver Metropolitan Area, Colorado, 2016–2017
  - Date: 1/1/17
  - Location: Denver metropolitan area
  - State: Colorado
  - CBSA Code: 19740
  - CBSA Title: Denver-Aurora-Lakewood, CO
- Notes from the Field: Mumps Outbreak — Alaska, May 2017–July 2018
  - Date: 5/1/17
  - Location: Anchorage
  - State: Alaska
  - CBSA Code: 11260
  - CBSA Title: Anchorage, AK
- Notes from the Field: Mumps Outbreak Associated with Cheerleading Competitions - North Texas, December 2016-February 2017
  - Date: 12/6/16
  - Location: Collin County, Texas
  - State: Texas
  - CBSA Code: 19100
  - CBSA Title: Dallas-Fort Worth-Arlington, TX
- Notes from the Field: Use of Social Media as a Communication Tool During a Mumps Outbreak — New York City, 2015
  - Date: 8/16/15
  - Location: Rockaways neighborhood of Queens
  - State: New York
  - CBSA Code: 35620
  - CBSA Title: New York-Newark-Jersey City, NY-NJ-PA

Varicella

- Notes from the Field: Varicella Outbreak Associated with Riding on a School Bus — Muskegon County, Michigan, 2015
  - Date: 12/3/15
  - Location: Muskegon County, Michigan
  - State: Michigan
  - CBSA Code: 24340
  - CBSA Title: Grand Rapids-Wyoming, MI

*Key Additional Results*

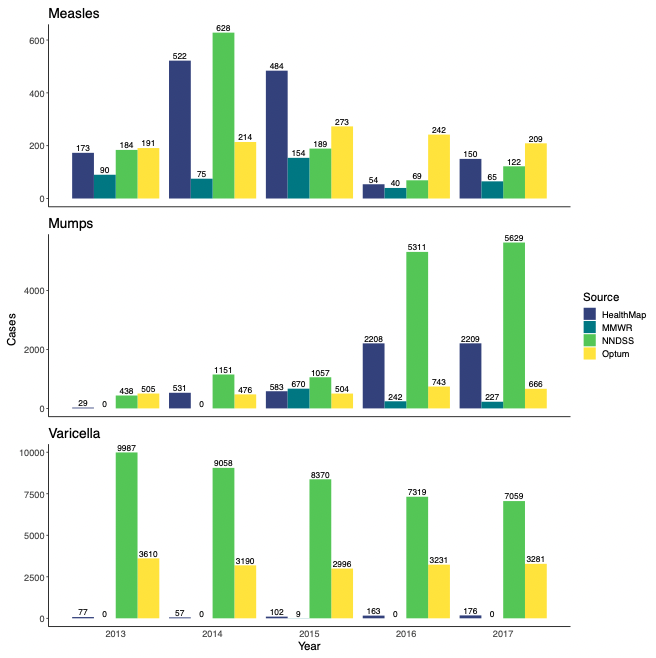

Supplement Figure 1. Unstandardized national-level case counts

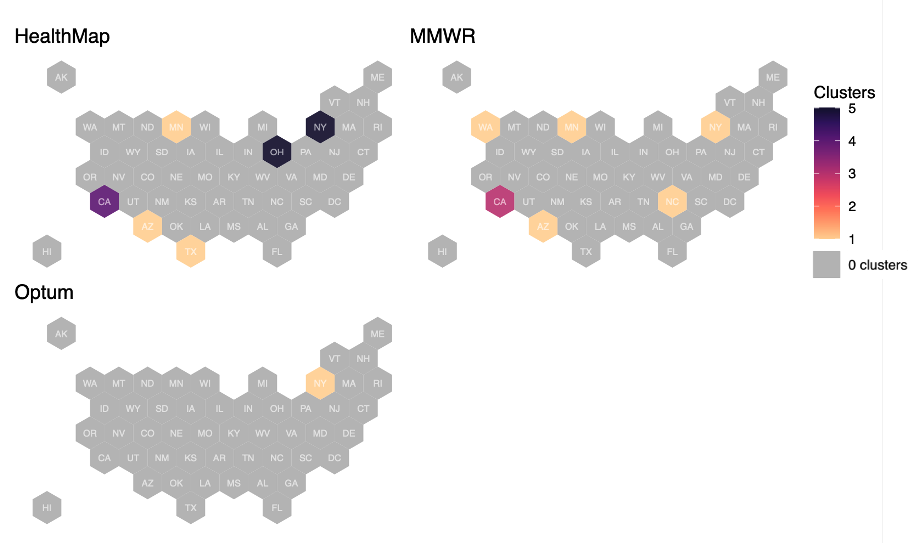

Supplement Figure 2. State-level measles clusters (2013–2017)

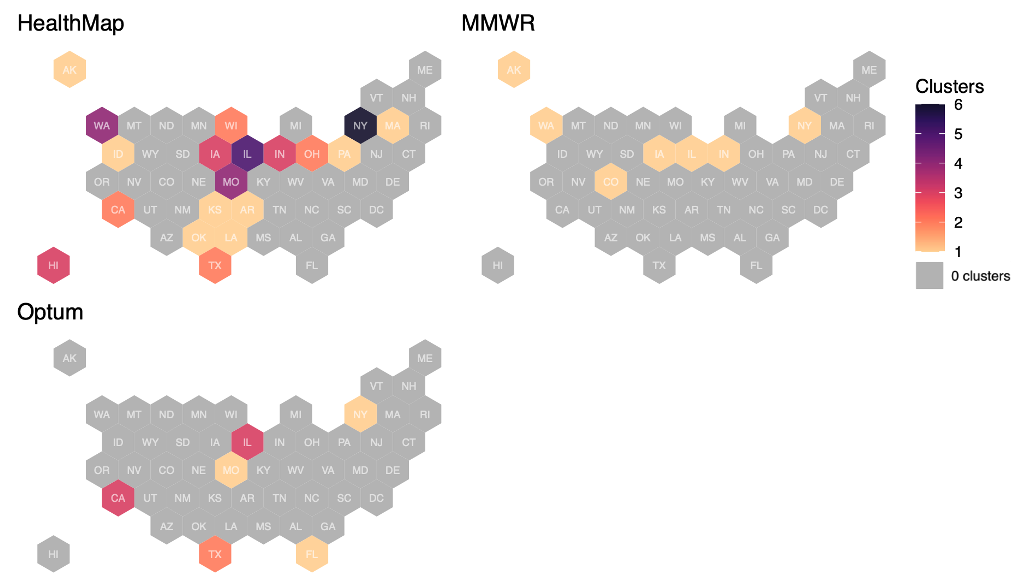

Supplement Figure 3. State-level mumps clusters (2013–2017)

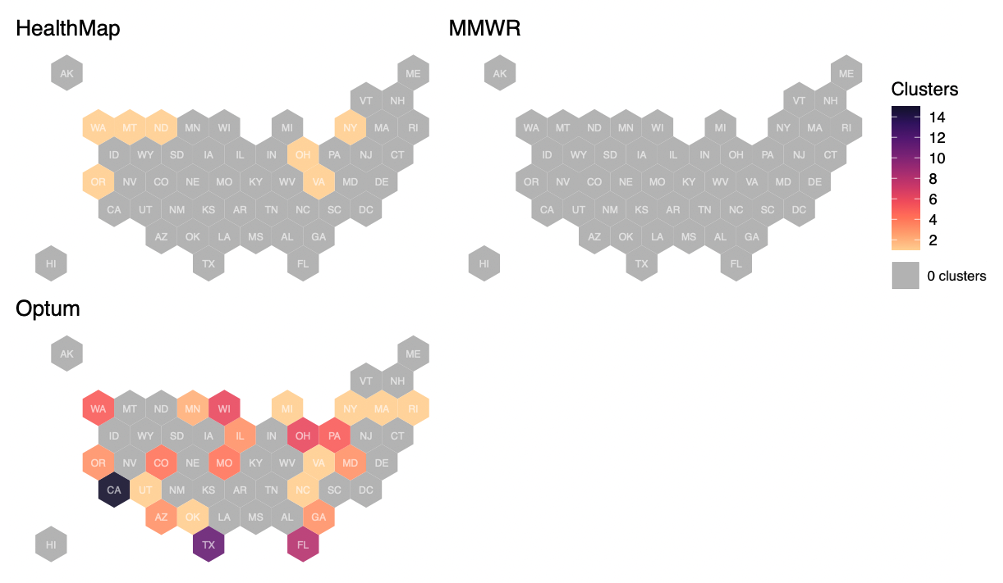
Supplement Figure 4. State-level varicella clusters (2013–2017)
